# NeuropsychBrainAge: a biomarker for conversion from mild cognitive impairment to Alzheimer’s disease

**DOI:** 10.1101/2022.11.29.22282870

**Authors:** Jorge Garcia Condado, Jesus M. Cortes, Alzheimer’s Disease Neuroimaging Initiative

## Abstract

**Background:** BrainAge models based on neuroimaging data have shown good accuracy for diagnostic classification. However, they have replicability issues due to site and patient variability intrinsic to neuroimaging techniques. We aimed to develop a BrainAge model trained on neuropsychological tests to identify a biomarker to distinguish stable mild cognitive impairment (sMCI) from progressive mild cognitive impairment (pMCI) to Alzheimer’s disease (AD).

**Methods:** Using a linear regressor, a BrainAge model was trained on healthy controls (CN) based on neuropsychological tests. The model was applied to sMCI and pMCI subjects to obtain predicted ages. The BrainAge delta, the predicted age minus the chronological age, was used as a biomarker to distinguish between sMCI and pMCI. We compared the model to one trained on neuroimaging features.

**Findings:** The AUC of the ROC curve for differentiating sMCI from pMCI was 0.91. It greatly outperforms the model trained on neuroimaging features which only obtains an AUC of 0.681. The AUC achieved is at par with the State-of-the-Art BrainAge models that use Deep Learning. The BrainAge delta was correlated with the time to conversion, the time taken for a pMCI subject to convert to AD.

**Interpretation:** We suggest that the BrainAge delta trained only with neuropsychological tests is a good biomarker to distinguish between sMCI and pMCI. This opens up the possibility to study other neurological and psychiatric disorders using this technique but with different neuropsychological tests.

**Funding:** A full list of funding bodies that supported this study can be found in the Acknowledgments section.

**Research in Context:** *Evidence before this study:* A major application of recent neuroimaging BrainAge models has been demonstrating its value in diagnostic classification. In spite of the good performance, most models based on neuroimaging data have limitations in real data as the distribution between sites can be different from training cohorts. They can also suffer from lack of specificity to a disease, for those based on BrainAge deltas trained on healthy controls or insufficient training data, for those trained to directly identify a specific disease. We develop a BrainAge model trained on neuropsychological tests used in Alzheimer’s disease research to identify a biomarker to distinguish sMCI from pMCI subjects. We propose a model that is trained on healthy controls for which there is more data to then reliably distinguish sMCI from pMCI subjects.

*Added value of this study:* This is the first study to use a BrainAge model based on neuropsychological test features to study Alzheimer’s disease. We suggest the NeuropsychBrainAge delta, which measure the difference between the model predicted age of the subject trained on healthy controls and the chronological age of the subject, as a biomarker of Alzheimer’s Disease. The NeuropsychBrainAge delta could differentiate between sMCI and pMCI. Moreover, we also show that the proposed biomarker is correlated with the time to conversion, the time taken for a pMCI subject to convert to Alzheimer’s Disease.

*Implications of all the available evidence:* Our approach could be used for the identification of patients with mild cognitive impairment at risk of developing Alzheimer’s disease. The NeuropsychBrainAge delta can also be used as a quantitative marker to measure disease severity due to its correlation with time to conversion. This study shows that using healthy controls for which there is more data but using features specific to a disease such as neuropsychological test can lead to reliable BrainAge models to identify specific neurological and psychiatric disorders.

## 1. Introduction

Recent advances in ageing modelling have been aided by the use of machine learning and deep learning to create BrainAge models [1–4] based on neuroimaging data. BrainAge models have rapidly been applied to the medical fields to identify neurological disorders, such as mild cognitive impairment (MCI) and Alzheimer’s disease (AD) [5–11], traumatic brain injury [12,13] and multiple sclerosis [14,15] and also psychiatric disorders, such as schizophrenia [16–19] and bipolar disorder [19,20]. There are two types of models used in classification tasks. Those that use the difference between subject’s predicted age trained on healthy controls (CN) and subject’s chronological age, BrainAge delta, as a biomarker for classification [1], and those that modify a deep learning model originally trained on BrainAge prediction and retrain the network on a classification task to distinguish CN from patients [4]. The first type of model suffers from a lack of specificity for a given disease and the BrainAge delta seems to vary considerably between studies and models [3]. The second type of models does not have a specificity problem, but may suffer from the lack of enough training data for the patient subjects. One of the critical limitations of using neuroimaging is the variability intrinsic to this type of imaging across sites. If analyses are not carried out appropriately, site effects can dominate and make the models unusable. This poses a great challenge when thinking about bringing these methods to a clinical setting. An alternative is to train these types of models with other less site-dependent features, such as neuropsychological tests.

We aimed to develop a BrainAge model trained on neuropsychological features that can be used to identify a biomarker of MCI to AD conversion. We used the data consisting only of cognitive normal subjects for training. The output of the model, defined as NeuropsychBrainAge delta in this study, represents the difference between the subject’s predicted age by the model and the subject’s chronological age. To show the applicability of this model to a neurological disorder in the clinical setting, we applied the model to a cohort of subjects with MCI, of whom some remained stable and others progressed to AD. The proposed biomarker is capable of distinguishing with good accuracy between stable MCI (sMCI), those who remain MCI, and progressive MCI (pMCI), those who converted to AD. There have been BrainAge models based on neuropsychological features to study cognitive age [21] and to predict age directly from behavioural tests [22]. However, to the authors’ knowledge, there are no previous studies to use these tests to predict age and then use the difference between predicted age and chronological age as a biomarker.

## 2. Methods

### 2.1 Subjects

Data used in the preparation of this article were obtained from the Alzheimer’s Disease Neuroimaging Initiative (ADNI) database (adni.loni.usc.edu) [23]. The ADNI was launched in 2003 as a public-private partnership, led by Principal Investigator Michael W. Weiner, MD. The primary goal of ADNI has been to test whether serial magnetic resonance imaging (MRI), positron emission tomography (PET), other biological markers, and clinical and neuropsychological assessment can be combined to measure the progression of mild cognitive impairment (MCI) and early Alzheimer’s disease (AD). For up-to-date information, see www.adni-info.org.

All subjects in the ADNI2 and ADNI3 phases who had an initial visit T1-weighted imaging and neuropsychological evaluation were extracted from the ADNI database. This included healthy controls (CN), mild cognitive impairment (MCI) and Alzheimer’s disease (AD) subjects. The dataset statistics is summarized in Table 1. Using longitudinal data, we identified conversors: those who converted from CN to MCI (N=64), those who converted from MCI to AD (N=152) and those who converted from CN to AD (N=7).

**Table 1.** Dataset Distribution

| Category | CN | MCI | AD |
| --- | --- | --- | --- |
| Nº Subjects | 629 | 635 | 208 |
| Age distribution (Years) | 72.1±6.8 | 72.5±7.8 | 74.7±8.2 |
| Nº Male / Nº Female | 265 / 364 | 356 / 279 | 121/87 |

A second dataset was created to study the effect of homogenisation on the results. For this dataset only sites with more than 10 CN subjects were used and which had at least 1 MCI and 1 AD subject. The CN and MCI cohorts were also resampled to ensure they have the same age distribution as the AD cohort.

The model was tested in the task of distinguishing between stable MCI (sMCI) subjects, those who remained MCI, and progressive MCI (pMCI) subjects, those who converted to AD. As it is impossible to know whether or not a sMCI subject will convert to AD at a future date, sMCI patients were considered stable if they remained with a diagnosis of MCI after 3 years from the initial visit. Similarly, only subjects with pMCI who converted to AD within 3 years from the initial visit were used. As there are more sMCI subjects, these were randomly sampled to obtain a balanced dataset containing equal amounts of sMCI (N=98) and pMCI (N=98) subjects.

### 2.2 Features

Models were built using two different types of features: structural brain features extracted from T1-weighted images and neuropsychological features. The first set of features were used to build a reference model so that we can compare our second model.

T1-weighted images from each subject’s first baseline visit were processed to obtain volumes of different brain structures. SIENAX [24], part of FSL [25] was used to obtain grey matter volume, white matter volume, cerebrospinal fluid volume, and peripheral grey matter volume as well as a volume scale value. FIRST [26] was used to segment and calculate the volumes of the thalamus, caudate, putamen, palidum, brainstem, hippocampus, amygdala and accumbens. The volume scale value was used to control for differences in brain size. For the dataset used for more powerful homogenisation, PyCombat [27] was used to homogenise taking into account the volume scale value, gender, and phase in which the patient was recruited. 12 features were used in total.

Neuropsychological assessments from each subject’s first baseline visit were used as features for the second model. This consisted of scores from standard neuropsychological tests: MMSE [28], ADAS [29], FAQ [30] and MOCA [31], as well as two metrics generated in the ADNI study: ADNI Memory score [32] and ADNI Executive Function [33]. 6 features were used in total.

### 2.3 Model

A linear regressor was trained in a supervised fashion (code can be found in https://github.com/JGarciaCondado/ADNIBrainAge). The task at hand is to use the features extracted from each subject at the initial visit to predict the age of the subject at the initial visit (Figure 1a). The model is trained using only CN subjects. For training 85% of all CN subjects are used and 15% are used for testing. The features are normalized by subtracting the training mean and dividing by the training standard deviation. The model can be summarized below:

**Figure 1.**
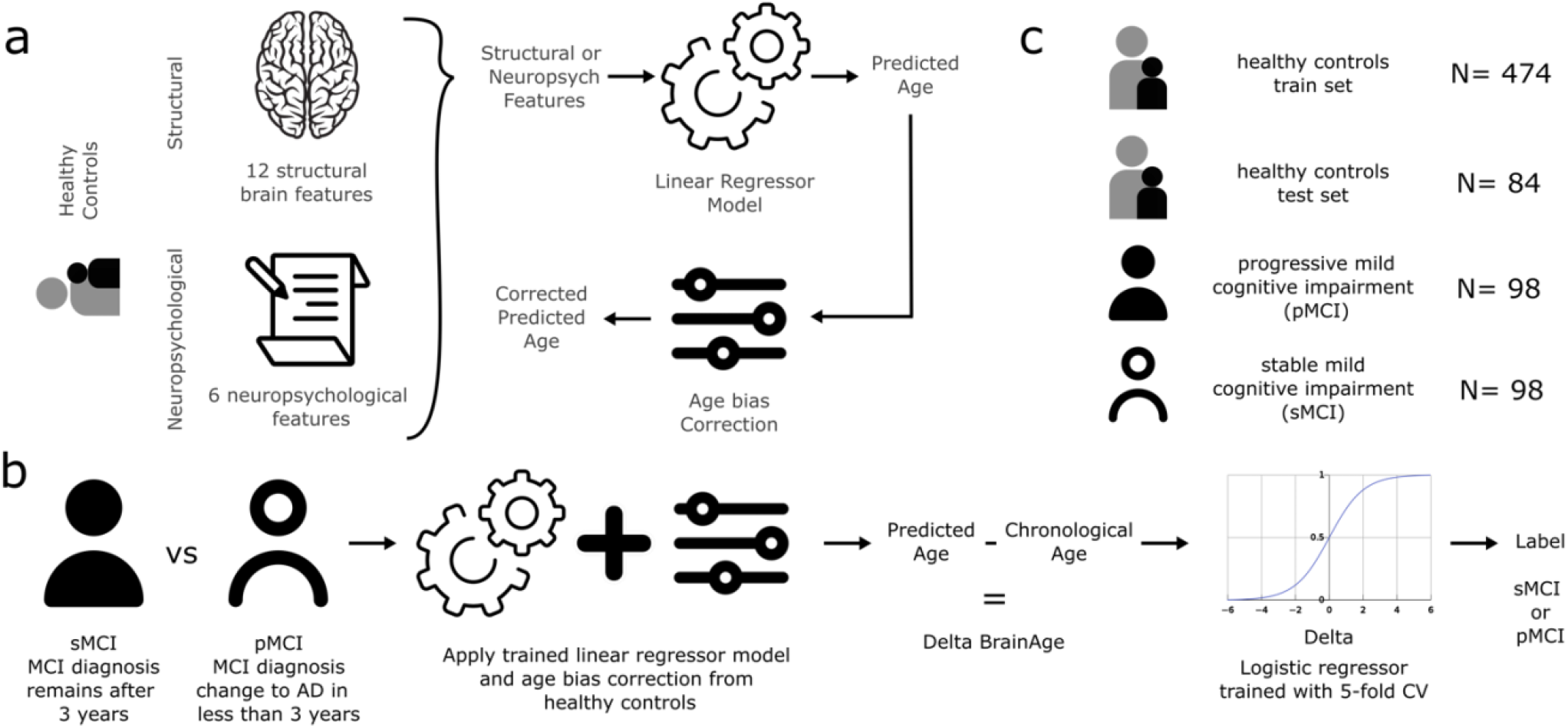
Overview of the BrainAge model and classification task. **a)** Training of the BrainAge model on healthy controls with input data consisting of structural features or neuropsychological features. A total of 12 structural brain features were used, consisting of volume measured in mm^3^ for: white matter, grey matter, peripheral grey matter, cerebrospinal fluid, thalamus, caudate, putamen, pallidum, hippocampus, amygdala, accumbens and brainstem. A total of 6 neuropsychological features were used: MMSE, ADAS, FAQ, MoCA, ADNI Memory and ADNI Executive Function. After training the linear regressor on the healthy controls age estimation task, an age bias correction was applied to deal with the inherent bias of regression to the mean problem. **b)** Description of the classification task between stable mild cognitive impairment (sMCI) and progressive mild cognitive impairment (pMCI). First, features were extracted for each subject as with healthy controls. Then, using either neuropsychological features or structural features, the trained model and bias correction were applied to obtain a predicted age. The BrainAge delta was calculated by subtracting the chronological age from the predicted age. This delta was then used as an input to a logistic regressor to determine a threshold for labelling using a 5-fold CV scheme. **c)** Number of subjects used for training with healthy controls, the number of subjects used to test the performance of BrainAge models on unseen healthy controls, and number of sMCI and pMCI used in the classification task.

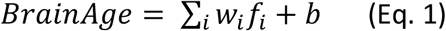

were f_i_ is the value of each normalized feature, w_i_ the weight of each feature and b the intercept. The weights and intercept are chosen to minimize the mean square error between the predicted age (Brain Age) and the subject’s age at the initial visit (Chronological Age, Ω):

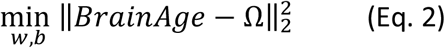

There is a bias in the model as younger controls tend to be given higher ages and older controls are given lower ages than they are as it is a regression to the mean problem. It can be thought of in terms of our best estimate for a subject which we know nothing about, being the mean, so younger patients tend to be given higher ages and older patients lower ages. This can be fixed by adjusting the predicted BrainAge taking into account the actual age of the training subjects to correct for this bias by first fitting the following model [34]:

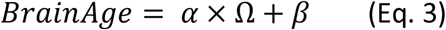

The coefficients *α* and *β* represent the slope and intercept, which are then used to correct the predictions in a test set using:

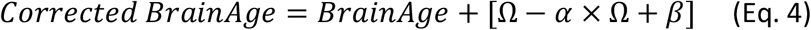

### 2.4 Biomarker

The model is trained on CN subjects only, so it is then applied to our test CN subjects, MCI subjects and AD subjects to predict their BrainAges. Our biomarker to differentiate our subjects is the BrainAge delta, defined as:

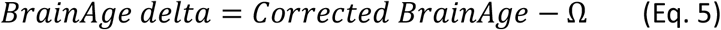

Notice that by using the corrected BrainAge, the BrainAge delta will not correlate with age but will be related to our variable of interest, the cohort each subject belongs to.

Finally, to assess the potential of BrainAge delta to differentiate between sMCI and pMCI subjects, a logistic regressor is trained using a 5-fold cross validation strategy on our task dataset (Figure 1b). Four logistic regressors are trained with the following inputs: one using StructBrainAge delta (delta found using the model trained using structural imaging features), one using NeuropsychBrainAge delta (delta found using the model trained using neuropsychological features), one using the sum of both deltas, and finally one using both deltas independently. Moreover, we also follow a different strategy of training logistic regressors but directly on the features (using only structural features, using neuropsychological features, and using all features) to compare to the logistic regressors trained on BrainAge deltas. In total 7 different logistic regressors were trained.

## 3. Results

### 3.1 Correlation between features and age

As a first initial assessment of the suitability of the proposed features for predicting age Pearson correlation coefficients were calculated between each feature and age of the CN subjects. Correlations between structural features and age is shown in Figure 2a, ranging from c_min_=0.12 (brainstem, p<0.02) to c_max_=0.55 (grey matter, p<3*×10*^*-39*^). As noted in previous research [35], there is a strong linear correlation between certain brain structures and population ageing above 50 years. Correlations between neuropsychological features and age is shown in Figure 2b, ranging from c_min_=0.08 (FAQ, p<0.10) to c_max_=0.35 (grey matter, p<7*×10*^*-15*^). These have a weaker correlation yet there is still one.

**Figure 2.**
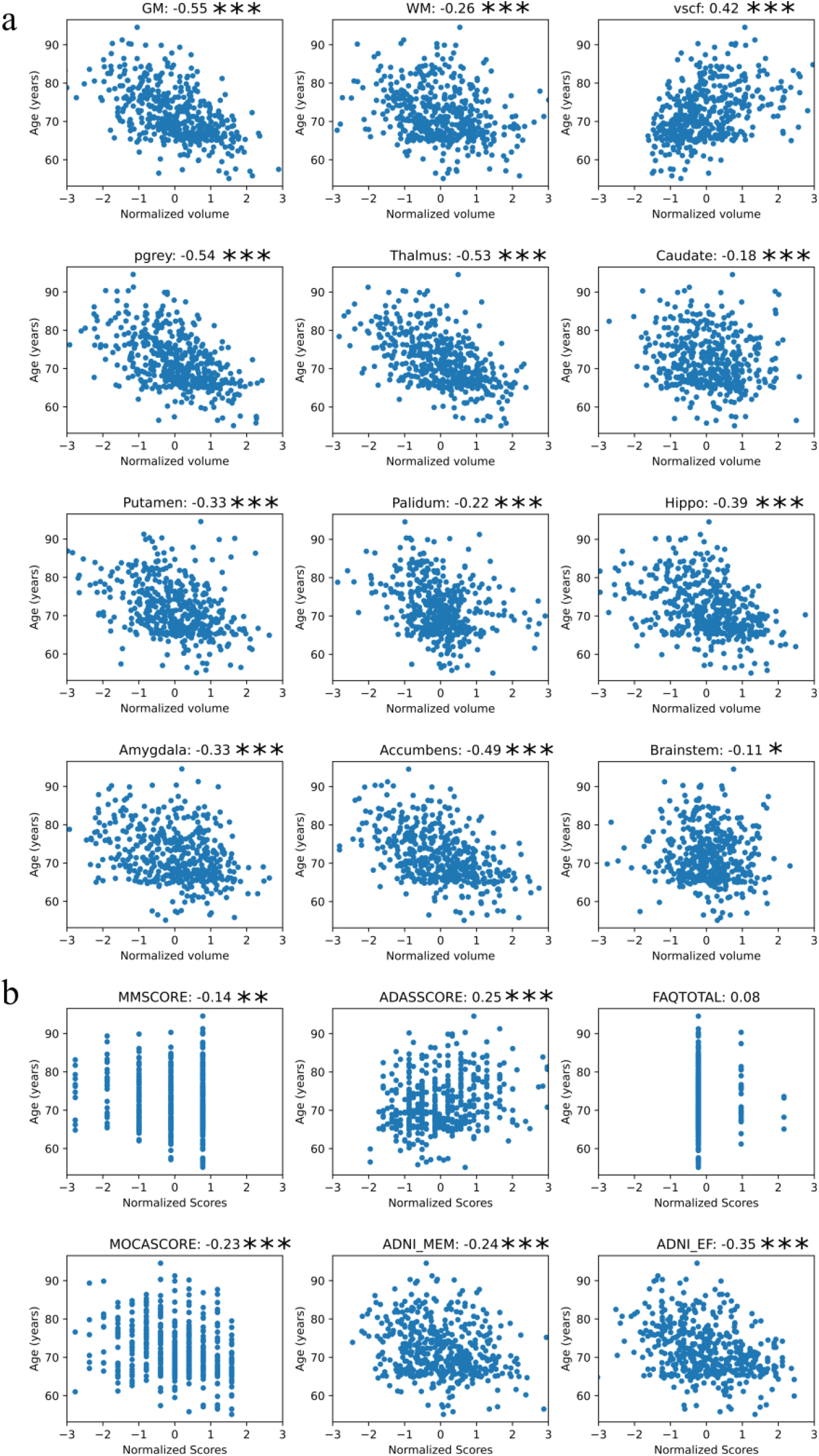
Correlation between BrainAge model features and age in healthy controls (CN) subjects resulting from **a)** Structural imaging features and **b)** Neuropsychological test features. The title of each graph describes the precise feature and its value of the Pearson’s correlation coefficient with age. *p < 0.05, **p<0.01, ***p < 0.001

### 3.2 A model trained only on healthy controls

A BrainAge model was trained on CN subjects on two set of features (neuroimaging and neuropsychological features) and tested on a different set of CN subjects. The results in Table 2 showcase the mean absolute error on the test set before and after correction for age bias.

**Table 2.**
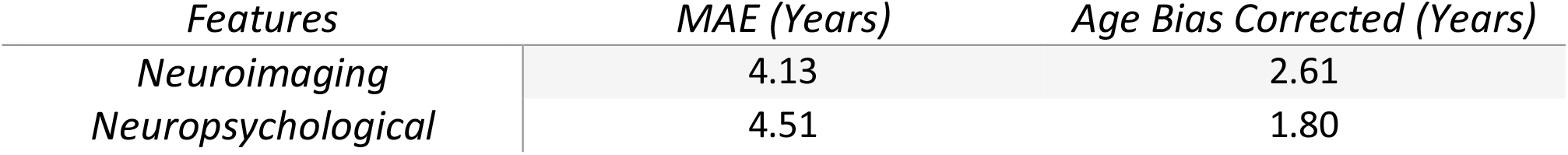
MAE of BrainAge models on test dataset of healthy controls without age bias correction and with age bias correction

### 3.3 Differences in BrainAge deltas between cohorts

We applied both BrainAge models, one trained on neuroimaging features and another trained on neuropsychological features, after age bias correction to all cohorts: the test set CN, MCI and AD subjects. Then we calculated the BrainAge delta for each subject. BrainAge delta was significantly higher between CN and MCI for both neuroimaging (−0.32±3.28 vs 1.92±3.97, *p<1×10*^*-7*^) and neuropsychology (−0.21±2.19 vs 4.42±3.95, *p<1×10*^*-26*^), as well as between MCI and AD subjects, for neuroimaging (1.92±3.97 vs 4.98±4.50, *p<1×10*^*-17*^) and neuropsychology (4.42±3.95 vs 16.07±6.38, *p<1×10*^*-83*^). The differences are shown in Figure 3a.

**Figure 3.**
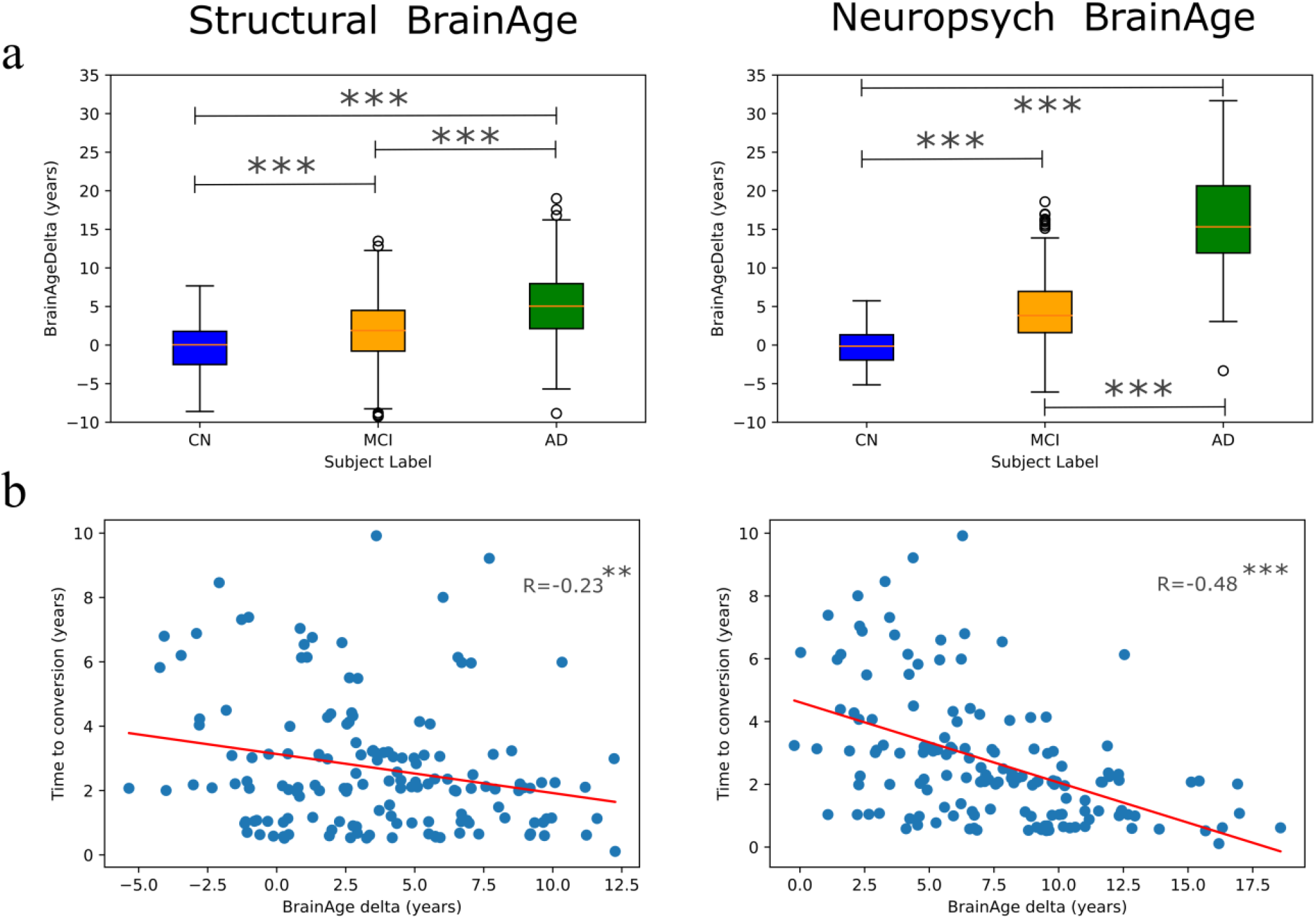
BrainAge delta for each subject group and its relation to conversion time for progressive mild cognitive impairment subjects. Structural BrainAge refers to the model trained on neuroimaging features and Neuropsych BrainAge refers to the model trained on neuropsychological features. **a)** BrainAge delta for each cohort, healthy control (CN), mild cognitive impairment (MCI) and Alzheimer’s disease. **b)** Time to conversion for subjects with progressive mild cognitive impairment (pMCI) as a function of BrainAge delta, where R indicates the Pearson correlation coefficient. Time to conversion is defined as the time between the subject’s first baseline visit and the visit at which the subject is labelled with Alzheimer’s disease (AD). *p < 0.05, **p < 0.01, ***p<0.001

We next addressed the BrainAge delta of the pMCI cohort, those labelled as MCI at the baseline visit but who would progress to AD during follow-ups. There was a correlation between the BrainAge delta and the conversion time, the number of years until the subject was labelled AD. Pearson’s correlation between conversion time and BrainAge delta was stronger when neuropsychological features were used (−0.48, *p<1×10*^*-10*^) as compared to neuroimaging features (−0.23, *p*<0.004), illustrated in Figure 3b.

### 3.4 Feasibility of application to identify sMCI vs pMCI subjects using BrainAge deltas

Next, we asked whether the Brain Age Delta could discriminate between sMCI subjects, those who remain MCI for at least 3 years after the initial visit from pMCI subjects, those who convert to AD within 3 years of the initial visit. A logistic regressor was trained on these BrainAge deltas using either the neuroimaging model (StructBrainAge delta), the neuropsychological model (NeuropsychBrainAge delta), the sum of both deltas, or each delta individually. The ROC curve for each can be seen in Figure 4. The best results were obtained when using the NeuropsychBrainAge deltas only as seen in Table 3.

**Table 3.** Results of different logistic regressors trained in a 5-fold CV scheme to distinguish between sMCI and pMCI subjects using different BrainAge deltas as inputs and possible combinations

| Model | AUC | Accuracy | Sensitivity | Specificity |
| --- | --- | --- | --- | --- |
| StructBrainAge delta | 0.68 [0.53-0.83] | 0.61 [0.51-0.72] | 0.63 [0.49-0.76] | 0.63 [0.47-0.80] |
| NeuropsychBra inAge delta | **0.91 [0.84-0.98]** | **0.85 [0.76-0.94]** | **0.87 [0.78-0.97]** | **0.87 [0.76-0.99]** |
| Addition | 0.86 [0.79-0.94] | 0.79 [0.69-0.89] | 0.81 [0.70-0.93] | 0.81 [0.66-0.96] |
| Multidomain | 0.90 [0.85-0.96] | **0.85 [0.77-0.93]** | 0.86 [0.77-0.95] | 0.85 [0.72-0.95] |

**Figure 4.**
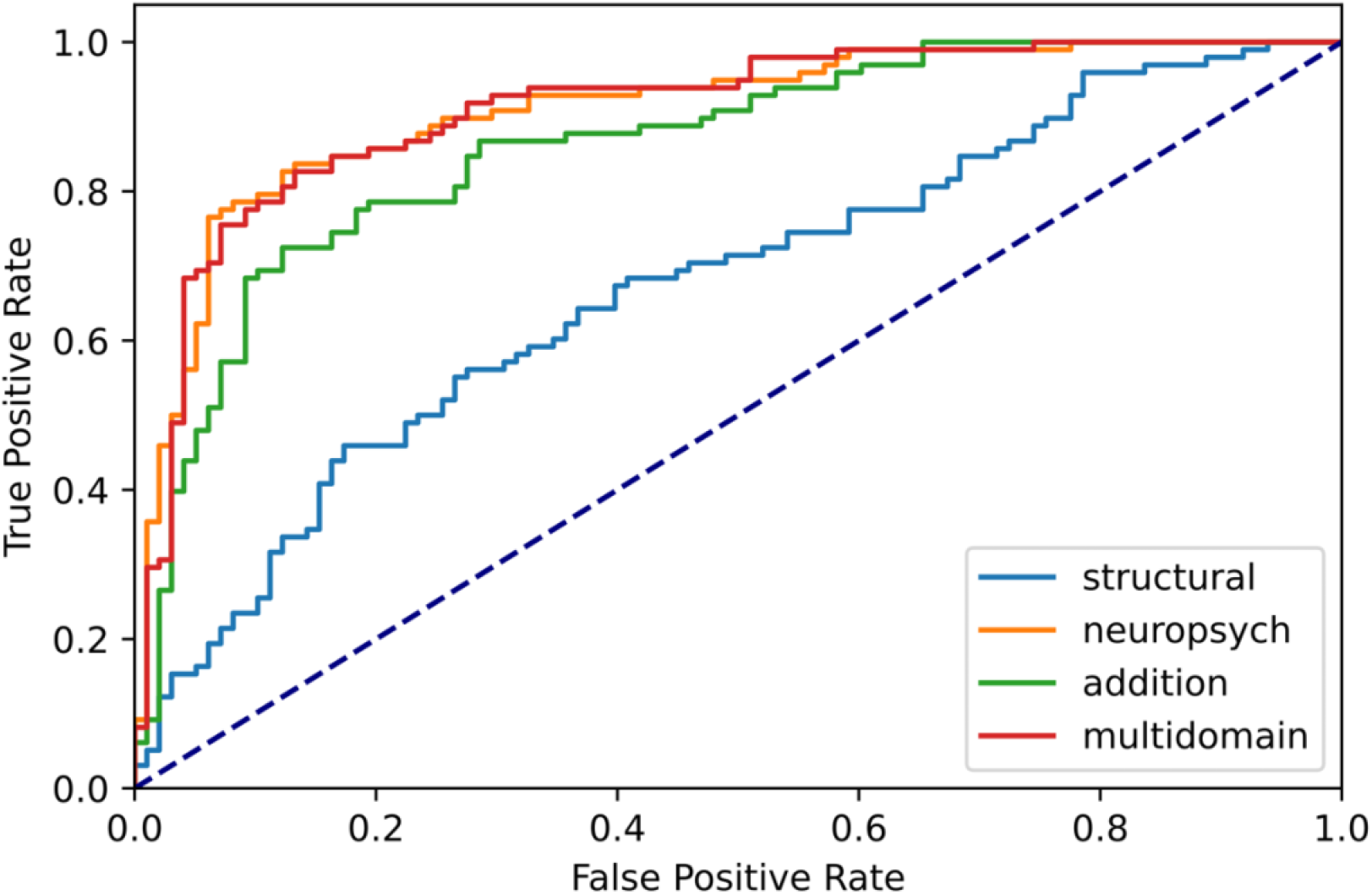
ROC curve for different logistic regressors models trained in a 5-fold CV scheme to discriminate between sMCI and pMCI subjects. (light blue) Structural refers to using the Structural BrainAge delta as the input to the logistic regressor. (orange) Neuropsych refers to using the Neuropsych BrainAge delta as the input to the logistic regressor. (green) Addition refers to using the sum of the Structural BrainAge delta and the Neuropsych BrainAge delta for each subject as a single input to the logistic regressor. (red) Multidomain refers to using Neuropsych BrainAge delta and Structural BrainAge delta as two separate inputs to the logistic regressor, hence in this logistic regressor there are two input features instead of one. (dashed) Represents the performance of a truly random classifier.

Further investigations were carried out to compare whether it would be better to use the features directly in a logistic regressor. For this purpose, logistic regressors to classify sMCI vs pMCI were trained using a 5-fold CV scheme directly on neuroimaging and neuropsychological features instead of BrainAge deltas as the input variables. The NeuropsychBrainAge delta-trained logistic regressor outperformed all other logistic regressors trained directly on neuroimaging or neuropsychological features, as shown in Table 4.

**Table 4.**
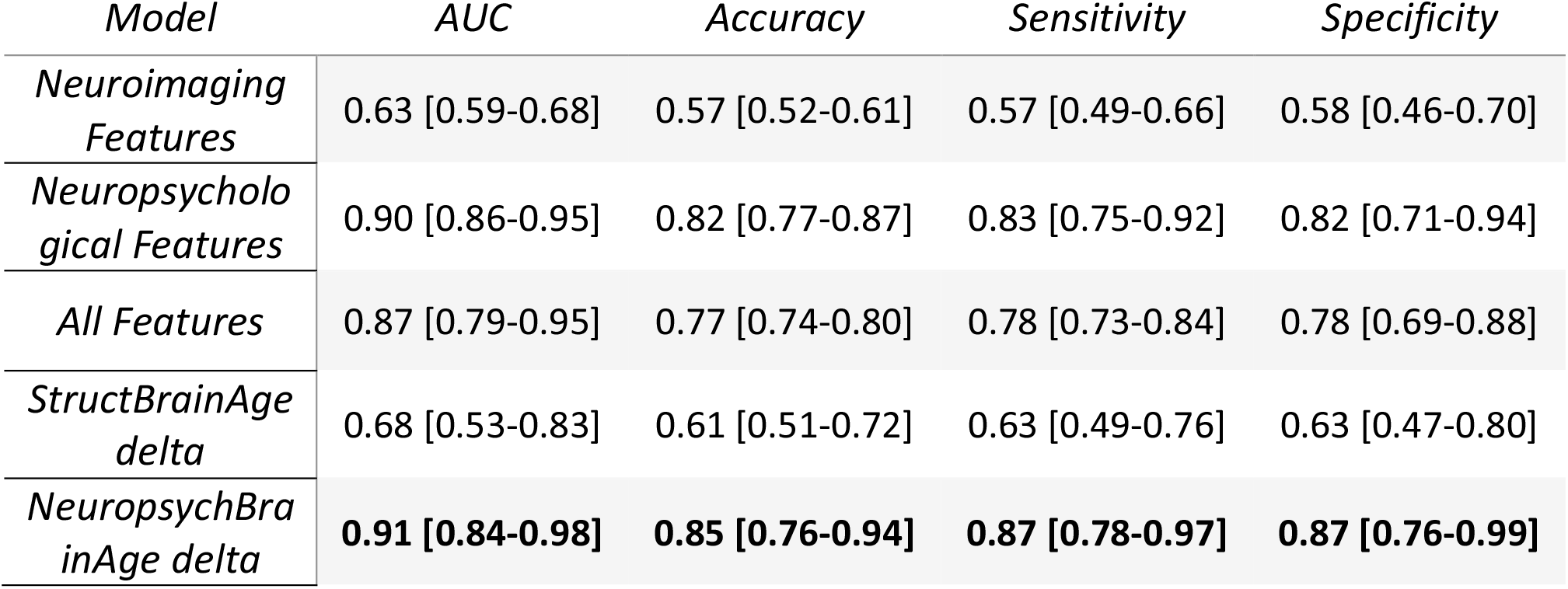
Results of different logistic regressors trained in a 5-fold CV scheme to distinguish between sMCI and pMCI subjects using individual features extracted as compared to BrainAge deltas

### 3.5 Effects of homogenisation of neuroimaging features

Lastly, we also tested whether the use of homogenisation techniques on neuroimaging features improved the performance of logistic regressors in the sMCI vs pMCI discrimination task. Neuroimaging homogenisation did improve performance (Table 5), but it was still lower than the performance achieved by NeuropsychBrainAge delta.

**Table 5.**
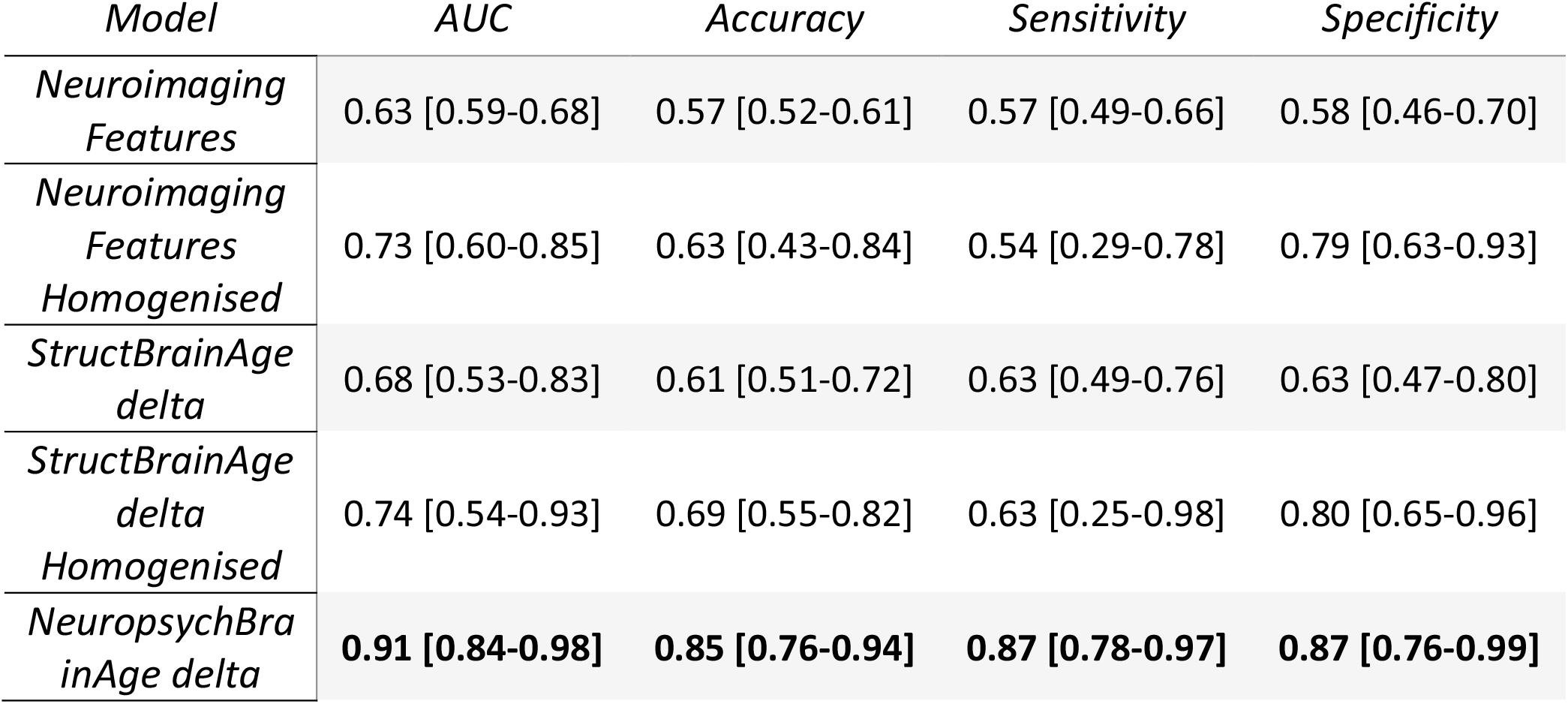
Performance of logistic regressors on neuroimaging features without and with homogenisation

## 4. Discussion

One of the key issues in the clinical application of BrainAge models is developing models that are reliable and reproducible. To achieve this, one strategy is to accurately track the trajectory of ageing in a normal healthy population. Deviations from this normal ageing trajectory can then indicate risks of developing certain conditions. The advantage of this type of modelling is twofold. The first is that by training the model on a healthy population and then tracking deviations from this, no assumptions are made about the condition under study that could bias the results. Second, it is much easier to collect data for healthy individuals and hence get larger datasets to train with. These models are then tested on the cohort with an underlying condition that we aim to identify to ensure the biomarker can correctly distinguish subjects in each cohort. Our BrainAge delta-trained logistic regressors were able to outperform logistic regressors trained directly on features because the deltas were extracted from a model trained on a larger dataset of healthy controls. BrainAge models were trained on healthy controls (N=474) and then the logistic regressors only had to find a threshold to divide the BrainAge deltas (a single feature) of sMCI (N=95) vs pMCI (N=95). On the other hand, logistic regressors trained directly on neuroimaging features had to combine information from multiple features (12 in the case of neuroimaging, 6 in the case of neuropsychological, or 18 in total) directly only on our task set of sMCI (N=95) vs pMCI (N=95) therefore leading to worse generalization due to lower number of datapoints.

The logistic regressor trained on NeuropsychBrainAge deltas was able to outperform all other models. It performs very similarly to using each BrainAge delta as separate inputs to the logistic regressor (multidomain approach), showing that all the information captured by StructBrainAge is already captured by NeuropsychBrainAge. In comparison to other State-of-the-Art models such as that original developed by Gaser et al. 2013 [1], based on structural imaging features and therefore similar to our study’s StructBrainAge, which achieved accuracies of 0.81, even lower than the 0.85 achieved by ours. It should be noted that the BrainAge model based on neuropsychological features performs worse in the task of predicting age, since its MAE in the test set for healthy controls is worse before bias correction than the model trained on neuroimaging data.

The large difference in accuracy between models trained in neuroimaging and neuropsychological tests, and after careful inspection of other models in the literature raises the question of whether the concept of BrainAge as a biomarker is robust and not strongly model dependent. Further studies are required to better understand whether there is a correlation between MAE before age bias correction and better performance on different classification tasks. There is a wide variety in reported BrainAge deltas between studies for similar cohorts. For example, AD subjects may have mean BrainAge deltas ranging from +5.35 years in one study to +10.70 in another [3]. This great variability indicates the need for further research on how to ensure that BrainAge-derived biomarkers are robust for clinical application across sites and subjects.

There might be concern in using neuropsychological tests to assess conversion, since they are used to assign the healthy control, MCI and AD labels in the first place. In ADNI, a selection of these tests are used in combination with cut-off points to assign those labels as well as other clinical assessments [23]. Various tests, such as CDR [36], are used to assign labels that we did not use in our study. However, the most important remark is that we trained our BrainAge model on healthy controls and then assessed the MCI conversion from baseline scores. There is no data leakage in terms of biases in the BrainAge deltas as the model is not trained on MCI subjects but on healthy controls. These tests have been used to assign subjects who belong to MCI at baseline, but we can predict future conversion to AD with those same scores without requiring future scores beyond the AD cut-off points. This is evidence to show that hard cut-off points are not the optimal tool for assigning labels as with hard cut-offs we label subjects into the same category (MCI) that have different pathological trajectories (sMCI vs pMCI). It is important to note that pMCIs are assigned as MCIs with the baseline scores because of the test cut-off scores and we can distinguish them from sMCI in the future with the baseline scores, but never using future assessment scores which make them their label to switch from MCI to AD. This shows that more information can be extracted from neuropsychological tests than is obvious by using cut-off points.

In this study we have shown that neuropsychological features combined with BrainAge modelling can yield a valuable biomarker to distinguish between sMCI and pMCI subjects. This biomarker also shows a strong correlation with conversion time, which is a sign of a robust biomarker as good biomarkers should have higher values with higher levels of severity, in this case, less time to conversion. Neuropsychological tests are better suited for clinical application as neuroimaging derived features suffer from a problem of greater variability between sites. Even when accounting for this with homogenisation techniques that make the pipeline more complex the neuropsychological based models still outperformed the neuroimaging ones. This comes to show that the use of BrainAge models combined with specific neuropsychological tests for a specific condition can provide valuable and accurate biomarkers.

## 5. Conclusion

We present a BrainAge model trained on neuropsychological tests able to discriminate stable MCI subjects from progressive MCI subjects. BrainAge models have the advantage of training on a larger cohort of healthy controls to measure deviations from the norm. By using features tightly linked to a specific condition, such as the neuropsychological tests for AD determination, we were able to achieve good performance on the classification tasks. Furthermore, we have shown that it is a robust biomarker because it was correlated with the conversion time from MCI patients to AD. We expect that our approach can be extended to other neurological and psychological disorders by applying the same models but with different neuropsychological tests specific to each condition.

## Data Availability

All data produced in the present study are available upon reasonable request to ADNI.

## Acknowledgments

The project that gave rise to these results received the support of a fellowship from “la Caixa” Foundation (ID 100010434). The fellowship code is LCF/BQ/DI21/11860030. J.M.C is funded by Ikerbasque: The Basque Foundation for Science and by the Department of Economic Development and Infrastructure of the Basque Country (Elkartek Program Grant KK-2021-00009).

Data collection and sharing for this project was funded by the Alzheimer’s Disease Neuroimaging Initiative (ADNI) (National Institutes of Health Grant U01 AG024904) and DOD ADNI (Department of Defense award number W81XWH-12-2-0012). ADNI is funded by the National Institute on Aging, the National Institute of Biomedical Imaging and Bioengineering, and through generous contributions from the following: AbbVie, Alzheimer’s Association; Alzheimer’s Drug Discovery Foundation; Araclon Biotech; BioClinica, Inc.; Biogen; Bristol-Myers Squibb Company; CereSpir, Inc.;Eisai Inc.; Elan Pharmaceuticals, Inc.; Eli Lilly and Company; EuroImmun; F. Hoffmann-La Roche Ltd. and its affiliated company Genentech, Inc.; Fujirebio; GE Healthcare; IXICO Ltd.; Janssen Alzheimer Immunotherapy Research & Development, LLC.; Johnson & Johnson Pharmaceutical Research & Development LLC.; Lumosity; Lundbeck; Merck & Co., Inc.; Meso Scale Diagnostics, LLC.; NeuroRx Research; Neurotrack Technologies; Novartis Pharmaceuticals Corporation; Pfizer Inc.; Piramal Imaging; Servier; Takeda Pharmaceutical Company; and Transition Therapeutics. The Canadian Institutes of Health Research is providing funds to support ADNI clinical sites in Canada. Private sector contributions are facilitated by the Foundation for the National Institutes of Health (www.fnih.org). The grantee organization is the Northern California Institute for Research and Education, and the study is coordinated by the Alzheimer’s Disease Cooperative Study at the University of California, San Diego. ADNI data are disseminated by the Laboratory for Neuro Imaging at the University of Southern California.

## Contributors

J.G.C. and J.M.C. designed the study. J.G.C. analyzed the data. J.G.C. and J.M.C. interpreted results and supported the analysis. All authors wrote and edited the manuscript. All authors approved the manuscript.

